# Impact of vaccination on the presence and severity of symptoms of hospitalised patients with an infection by the Omicron variant (B.1.1.529) of the SARS-CoV-2 (subvariant BA.1)

**DOI:** 10.1101/2022.10.23.22281414

**Authors:** Guillaume Beraud, Laura Bouetard, Rok Civljak, Jocelyn Michon, Necla Tulek, Sophie Lejeune, Romain Millot, Aurélie Garchet-Beaudron, Maeva Lefebvre, Petar Velikov, Benjamin Festou, Sophie Abgrall, Ivan Kresimir Lizatovic, Aurélie Baldolli, Huseyin Esmer, Sophie Blanchi, Gabrielle Froidevaux, Nikol Kapincheva, Jean-François Faucher, Mario Duvnjak, Elçin Afşar, Luka Švitek, Saliha Yarimoglu, Rafet Yarimoglu, Cécile Janssen, Olivier Epaulard

## Abstract

**Objectives:** The emergence of SARS-CoV-2 variants raised questions over the extent to which vaccines designed in 2020 have remained effective. We aimed to assess whether vaccine status was associated with the severity of Omicron SARS-CoV-2 infection in hospitalised patients.

**Methods:** We conducted an international, multicentric, retrospective study in 14 centres (Bulgaria, Croatia, France, Turkey). We collected data on patients hospitalised ≥24 hours between 01/12/2021 and 03/03/2022, with PCR-confirmed infection at a time of exclusive Omicron circulation, with hospitalisation related or not to the infection. Patients who had received prophylaxis by monoclonal antibodies were excluded. Patients were considered fully vaccinated if they had received at least 2 injections of either mRNA and/or ChAdOx1-S, or 1 injection of Ad26.CoV2-S vaccines.

**Results:** Among the 1215 patients (median [IQR] age 73.0 [57.0; 84.0]; 51.3% males), 746 (61.4%) were fully vaccinated. In multivariate analysis, being vaccinated was associated with lower 28-day mortality (RR=0.50 [0.32-0.77]), ICU admission (R=0.40 [0.26-0.62], and oxygen requirement (RR=0.34 [0.25-0.46]), independently of age and comorbidities. When co-analysing these Omicron patients with 948 Delta patients from a study we recently conducted, Omicron infection was associated with lower 28-day mortality (RR=0.53 [0.37-0.76]), ICU admission (R=0.19 [0.12-0.28], and oxygen requirements (RR=0.50 [0.38-0.67]), independently of age, comorbidities and vaccination status.

**Conclusions:** mRNA- and adenovirus-based vaccines have remained effective on severity of Omicron SARS-CoV-2 infection. Omicron is associated with a lower risk of severe forms, independently of vaccination and patient characteristics.

## Introduction

First identified in Botswana and South Africa in November 2021, the Omicron (B.1.1.529) variant of concern (VOC) of SARS-CoV-2 replaced the Delta (B.1.617.2) VOC globally, and became virtually the only circulating variant in most countries in December 2021 and January 2022. Between December 2021 and April 2022, it was responsible for the most massive waves so far of SARS-CoV-2 infection in Europe, North America, China and Japan since the beginning of the pandemic (ECDC 2022). The range of Omicron VOC waves is considered to be related to increased infectivity and to immune escape from neutralising antibodies resulting from previous SARS-CoV-2 infection, vaccination, or both (Syed et al. 2022; Cui et al. 2022).

In most European countries, the unprecedented numbers of SARS-CoV-2 infections observed during these Omicron waves were associated with a decreased case-fatality rate compared with the Delta VOC (Nyberg et al. 2022; Johnson et al. 2022; Auvigne et al. 2022). Whether related to an intrinsically lower risk of severe COVID-19 associated with Omicron infection, or to the high vaccine coverage in most of these countries, this VOC calls into question the impact of vaccination at both the individual and the populational level. Meanwhile, the picture provided by the aforementioned studies is incomplete, as most milder cases will not be admitted to hospital or even be diagnosed.

We aimed to determine the extent to which vaccination was still associated with protection against severe forms of Omicron infection, by assessing whether such forms differed among people hospitalised with SARS-CoV-2 infection when considering their vaccine status, as well as other risk factors. We also explored the influence of VOC by coupling our data with previously published data with the Delta variant, and using the same methodology.

## Methods

We carried out a retrospective, multicentre study across 14 hospital centres from four countries, namely Bulgaria (Sofia), Croatia (Olsijek, Zagreb), France (Annecy, Paris/Béclère, Caen, Grenoble, Le Mans, Limoges, Nantes, Poitiers) and Turkey (Ankara and Karaman). Hospitalised patients with a PCR-confirmed SARS-CoV-2 infection from December, 1st 2021 to March, 3rd 2022 (i.e., when the Omicron (B.1.1.529) variant [subvariant BA.1] was virtually the only circulating SARS-CoV-2 variant in these countries) were included chronologically regardless of the indication for admission, the objective being to include at least the first 50 to 100 patients per centre. Patients were considered as infected by the Omicron VOC either through mutation screening, or because it was the only variant circulating at that time in the area according to the virologist from each centre.

A patient was considered fully vaccinated if he/she had received at least either one injection of the Ad26.CoV2-S vaccine (Janssen), or two injections of the ChAdOx1-S vaccine (AstraZeneca) and/or RNA vaccines (tozinameran [Pfizer/BioNTech] and elasomeran [Moderna]), and with the last injection given at least 14 days before the date of PCR. Vaccinated and unvaccinated patients were included concurrently, according to order of hospitalisation in the participating centre.

Data were anonymously collected with a form similar to the one used in a recent study on SARS-CoV-2 Delta infection [1], providing similar variables (Supplementary material, Table 1 and 2), with the exception of antiviral treatments which were not available at the time data were collected for the Delta study. The ethics committee of the French-speaking Society of Infectious Diseases (SPILF) (IRB00011642) gave its approval for the study (N°2022-0101), which was declared to the French National Commission for Informatics and Liberties (CNIL MR004: n°2224742).

**Table 1:** Characteristics of patients with SARS-CoV-2 (Omicron variant) infection according to vaccinal status: Demographics, underlying diseases and presentation

|  | Vaccinated (N=746) | Unvaccinated (N=469) | p-value |
| --- | --- | --- | --- |
| Age (years) n (%) |  |  | 0.300 |
| <35y | 67 (9.0) | 54 (11.5) |  |
| 35-65y | 198 (26.5) | 128 (27.3) |  |
| >65y | 481 (64.5) | 287 (61.2) |  |
| Age (years) | 74.0 [57.3-85.0] | 71.0 [55.0-83.0] | 0.041 |
| Gender: men n (%) | 406 (54.4) | 217 (46.3) | 0.007 |
| Body Mass Index (kg/m<sup>2</sup>) | 25.9 [22.2-29.4]<sub>(n=391)</sub> | 26.1 [22.6-29.2]<sub>(n=232)</sub> | 0.670 |
| Overweight (BMI>25) n (%) | 221 (56.5)<sub>(n=391)</sub> | 134 (57.8)<sub>(n=232)</sub> | 0.828 |
| Pre-existing comorbidities n (%) |  |  |  |
| Cardiac failure | 121 (23.6)<sub>(n=512)</sub> | 65 (18.3)<sub>(n=355)</sub> | 0.073 |
| Respiratory disease | 103 (13.8) | 47 (10.0) | 0.062 |
| Oxygen at home | 15 (2.9)<sub>(n=514)</sub> | 3 (0.8)<sub>(n=354)</sub> | 0.063 |
| Kidney failure (GFR<30) | 88 (12.0)<sub>(n=731)</sub> | 31 (6.8)<sub>(n=459)</sub> | 0.004 |
| Diabetes | 184 (24.7) | 106 (22.7) | 0.452 |
| Hypertension | 261 (50.8)<sub>(n=514)</sub> | 183 (51.5)<sub>(n=355)</sub> | 0.877 |
| Solid cancer < 3 months | 22 (3.0)<sub>(n=745)</sub> | 9 (1.9) | 0.351 |
| Hematologic malignancy | 42 (5.6) | 10 (2.1)<sub>(n=468)</sub> | 0.003 |
| Immunodepression | 77 (15.0)<sub>(n=514)</sub> | 33 (9.3)<sub>(n=355)</sub> | 0.013 |
| At least one comorbidity among the above | 523 (70.1) | 330 (70.4) | 0.949 |
| Number of comorbidities | 1.0 [0.0; 2.0] | 1.0 [0.0; 2.0] | 0.201 |
| Tobacco, active or discontinued < 3 years, n (%) | 44 (11.9)<sub>(n=370)</sub> | 38 (15.4)<sub>(n=247)</sub> | 0.258 |
| Previous SARS-CoV-2 infection n (%) | 12 (1.9)<sub>(n=633)</sub> | 10 (2.4)<sub>(n=415)</sub> | 0.661 |
| Time from a previous infection (days) | 319.0 [43.0; 344.0]<sub>(n=9)</sub> | 215 [97.3; 389.5]<sub>(n=10)</sub> | 0.497 |
| Time from last vaccine dose (days) | 113.0 [64.0; 178.2] | - | - |
| Admission for COVID-19 | 359 (48.7)<sub>(n=737)</sub> | 317 (69.5)<sub>(n=456)</sub> | <0.001 |
| Ct value (lowest value if many) | 21.4 [18.2; 27.0]<sub>(n=423)</sub> | 22.1 [19.0; 27.3]<sub>(n=273)</sub> | 0.125 |
| CRP (highest value) (mg/L) | 58.0 [21.5; 117.5]<sub>(n=575)</sub> | 66.0 [26.0; 126.3]<sub>(n=380)</sub> | 0.110 |
| Proportion of lung involved on CT scan n (%) | n=179 | n=168 | 0.003 |
| 0-10% | 90 (50.3) | 51 (30.4) |  |
| 11-25% | 40 (22.3) | 55 (32.7) |  |
| 26-50% | 32 (17.9) | 41 (24.4) |  |
| 51-75% | 12 (6.7) | 18 (10.7) |  |
| >75% | 5 (2.8) | 3 (1.8) |  |

Quantitative variables are presented as median (1st quartile - 3rd quartile). When data were missing, or calculation is performed on a subgroup, the number of patients out of whom the statistic was calculated is mentioned alongside.

**Table 2:**
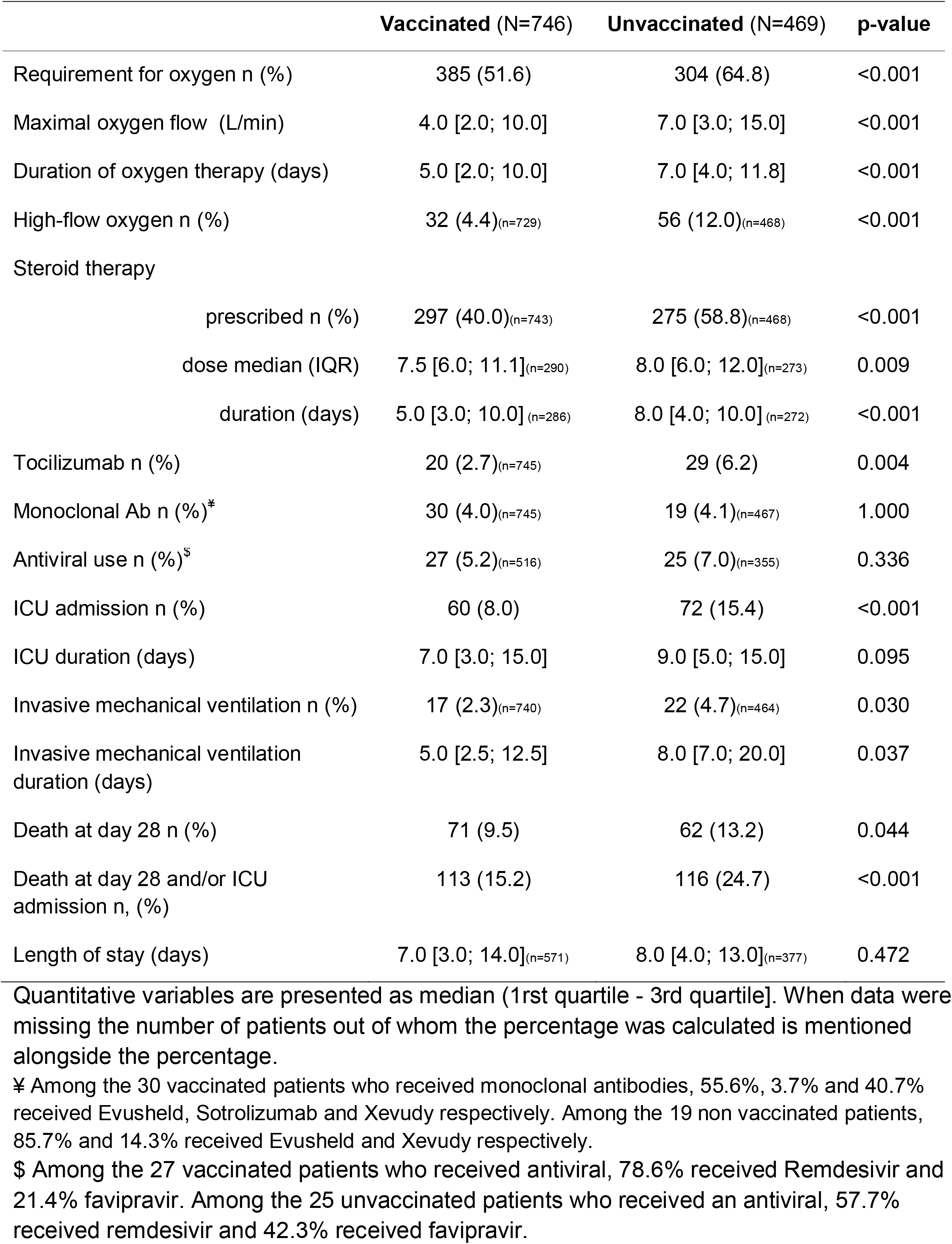
Characteristics of patients with SARS-CoV-2 (Omicron variant) infection according to vaccinal status: Management and outcomes during hospital stay

### Statistical analysis

The cohort population was described according to vaccination status with qualitative variables as counts (percentage) and frequency distributions compared with the Chi square test or Fisher’s exact test when appropriate. Continuous variables were expressed as median (1^rst^ quartile; 3^rd^ quartile) and differences were tested with the independent t-test for normally distributed variables or otherwise the Mann-Whitney U test. Factors associated with severe forms of infection defined by three outcomes (requirement for oxygen, ICU admission and death at day 28) were assessed. Univariate and multivariate logistic regressions based on general linear models were performed with a stepwise variable selection according to the Akaike Information Criterion (Akaike 1973). To assess the impact of comorbidities and vaccination, we selected all the potential risk factors for severe outcome presented in Table 3 for the multivariate model (Table 4 and Supplementary materials). Even though some data were missing at random, in many cases this was because they did not exist. As an example, missing CT-scan results could have been due to the fact that patients were too severe to receive a CT scan before high flow nasal oxygen or mechanical ventilation, or to the fact that the patient was only mildly ill and went back home without any oxygen requirement. To manage Missing data Not At Random (MNAR), multivariate analysis was initially performed only on the dataset with complete cases, after which the model was applied to the full dataset (Jakobsen et al. 2017). Results are presented based on the full dataset, as they were similar to the analysis restricted to the complete case dataset. In addition, a multivariate analysis was similarly carried out on the dataset of vaccinated patients only, so as to independently assess the weight of comorbidities among vaccinated patients, taking into account the influence of the time elapsed since the most recent injection. Interactions were systematically searched and mentioned if found. Finally, this cohort of Omicron-infected patients was merged with a cohort of Delta-infected patients recently published (Epaulard et al. 2022), and an analysis was performed taking into account the variant of concern.

**Table 3:** Association of patient characteristics with negative outcomes (bivariate logistic regression)

| Factors | Death at day 28 |  | ICU |  | Oxygen required |  |
| --- | --- | --- | --- | --- | --- | --- |
|  | OR (IC95%) | p-value | OR (IC95%) | p-value | OR (IC95%) | p-value |
| Age (years) | 1.05 [1.04; 1.07] | <0.001 | 1.00 [0.99; 1.00] | 0.306 | 1.03 [1.02; 1.04] | <0.001 |
| Gender: Male | 1.14 [0.79; 1.64] | 0.485 | 1.48 [1.03; 2.14] | 0.038 | 1.38 [1.10; 1.73] | 0.006 |
| Diabetes mellitus | 1.55 [1.04; 2.28] | 0.028 | 1.64 [1.10; 2.40] | 0.014 | 1.31 [1.00; 1.72] | 0.048 |
| Hypertension | 1.47 [0.97; 2.27] | 0.073 | 1.21 [0.80; 1.83] | 0.365 | 1.87 [1.43; 2.45] | <0.001 |
| Tobacco | 1.44 [0.66; 2.87] | 0.323 | 1.53 [0.67; 3.14] | 0.278 | 1.16 [0.72; 1.85] | 0.546 |
| BMI (kg/m<sup>2</sup>) | 0.94 [0.89; 0.99] | 0.037 | 1.05 [1.00; 1.09] | 0.029 | 1.07 [1.04; 1.10] | <0.001 |
| Overweight (BMI>25) | 0.54 [0.30; 0.97] | 0.040 | 1.44 [0.83; 2.59] | 0.207 | 1.71 [1.24; 2.36] | 0.001 |
| Kidney failure (GFR<30) | 2.86 [1.77; 4.53] | <0.001 | 2.11 [1.26; 3.42] | 0.003 | 1.81 [1.21; 2.75] | 0.005 |
| Respiratory disease | 1.67 [1.01; 2.66] | 0.036 | 1.40 [0.83; 2.27] | 0.189 | 3.06 [2.06; 4.64] | <0.001 |
| Oxygen at home | 0.97 [0.15; 3.48] | 0.968 | 1.48 [0.34; 4.59] | 0.539 | 15.04 [3.07; 271.60] | 0.009 |
| Cardiac failure | 1.71 [1.06; 2.69] | 0.024 | 1.39 [0.86; 2.19] | 0.167 | 1.58 [1.14; 2.22] | 0.007 |
| Solid cancer < 3 months | 5.54 [2.56; 11.57] | <0.001 | 0.88 [0.21; 2.51] | 0.829 | 1.06 [0.52; 2.23] | 0.874 |
| Hematologic malignancy | 2.29 [1.09; 4.42] | 0.019 | 0.49 [0.12; 1.36] | 0.236 | 1.34 [0.76; 2.43] | 0.314 |
| Immunosuppression | 1.64 [0.92; 2.80] | 0.082 | 1.28 [0.70; 2.22] | 0.397 | 1.23 [0.82; 1.85] | 0.318 |
| At least one comorbidity | 1.52 [1.00; 2.36] | 0.055 | 1.93 [1.25; 3.11] | 0.004 | 1.65 [1.29; 2.11] | <0.001 |
| Number of comorbidities | 1.26 [1.11; 1.42] | <0.001 | 1.20 [1.06; 1.36] | 0.003 | 1.20 [1.10; 1.30] | <0.001 |
| Previous SARS-CoV-2 infection | 0.40 [0.05; 2.98] | 0.368 | 0.42 [0.06; 3.17] | 0.402 | 0.81 [0.35; 1.90] | 0.630 |
| Time from last infection (days) <sup>£</sup> |  | 1.000 |  | 0.997 |  | 0.739 |
| Vaccination | 0.69 [0.48; 0.99] | 0.045 | 0.48 [0.33; 0.69] | <0.001 | 0.58 [0.46; 0.73] | <0.001 |
| Time from last vaccine injection (days) <sup>£</sup> |  | 0.289 |  | 0.117 |  | 0.104 |

£: For the time from last infection, and from last vaccine injection, the scarcity of data made the calculation of confidence intervals irrelevant.

**Table 4:** Factors associated with negative outcomes, with a multivariate logistic regression (best model according to AIC).

|  | Death on day 28 |  | ICU |  | Need for oxygen |  |
| --- | --- | --- | --- | --- | --- | --- |
|  | Best model | p-value | Best model | p-value | Best model | p-value |
| Age | 1.06 [1.04; 1.08] | <0.001 |  |  | 1.03 [1.02; 1.03] | <0.001 |
| Gender: Male |  |  | 1.25 [0.82; 1.90] | 0.304 | 1.37 [1.02; 1.83] | 0.036 |
| Diabetes |  |  | 1.51 [0.95; 2.36] | 0.076 |  |  |
| Kidney failure | 2.52 [1.44; 4.31] | 0.001 | 2.30 [1.29; 3.97] | 0.004 |  |  |
| Previous SARS-CoV-2 infection | 0.29 [0.02; 1.59] | 0.247 | 0.36 [0.02; 1.81] | 0.325 |  |  |
| Vaccination | 0.50 [0.32; 0.77] | 0.002 | 0.40 [0.26; 0.62] | <0.001 | 0.34 [0.25; 0.46] | <0.001 |
| Pulmonary diseases | 1.51 [0.86; 2.56] | 0.138 | 1.68 [0.96; 2.85] | 0.060 | 1.91 [1.19; 3.12] | 0.009 |
| O<sub>2</sub> at home |  |  |  |  | 8.41 [1.58; 156.23] | 0.044 |
| Solid cancer for < 3 months | 7.99 [2.97; 20.80] | <0.001 |  |  |  |  |
| Haematological cancer | 3.84 [1.67; 8.29] | 0.001 |  |  |  |  |
| Immunosuppressed |  |  |  |  | 1.58 [1.03; 2.46] | 0.038 |

Models were explored with a full model including the variable "gender", "Age", "Diabetes", "HTA", "KidneyFailure", "O2\_home", "CardiacFailure", "ImmunoSup", "previousSARS-CoV-2 infection", "Vaccination", "PulmDis", "SolidCancer3M", "HematoK", "OneComorb". "Tobacco", "BMI", "overweight" were not included due to more than 50% of missing data. No interactions were found.

All statistical analyses were performed with R version 4.1.2. Packages readxl, Amelia, xlsx, ggplot2, MASS, VIM were used.

## Results

We included 1215 hospitalised patients, among whom 746 (61.4%) were fully vaccinated and 469 (38.6%) were not vaccinated at all. Main characteristics of both groups are described in Table 1.

Among vaccinated patients with complete information on their vaccine history (n=674), most were vaccinated with Pfizer vaccines only (n=431, 63.9%) or a mix of the two mRNA vaccines (n=91, 13.5%) whereas a few had had a mix of mRNA and adenovirus vaccines (53, 7.9%), or only Moderna vaccines (n=52, 7.7%), AstraZeneca vaccines (n=23, 3.4%) or Janssen vaccines (n=15, 2.2%). Three hundred and ninety-five patients (53.7% of the vaccinated) had received more than 2 doses. The median time from the last vaccine dose was 113.0 days [IQR 64.0-178.2].

### Characteristics of the SARS-CoV-2 infection according to vaccine status

In bivariate analysis, vaccinated patients were slightly older, more frequently male, and more frequently presented with certain comorbidities (respiratory disease, hematologic malignancy, immunodepression), although they did not differ with unvaccinated patients in terms of median number of comorbidities.

Notwithstanding which, unvaccinated patients had more severe SARS-CoV-2 infections (Table 2): they were more frequently admitted because of COVID-19 instead of being admitted for another cause with an incident diagnosis of an asymptomatic SARS-CoV-2 infection; their lung involvement was more extended on CT scan; they more frequently required oxygen supplementation, at higher flow, and for a longer duration; they more frequently required high-flow oxygen therapy, and immunomodulatory therapy such as steroids or tocilizumab. The protective effect of vaccination predominated at an older age (Figure 1).

**Figure 1:**
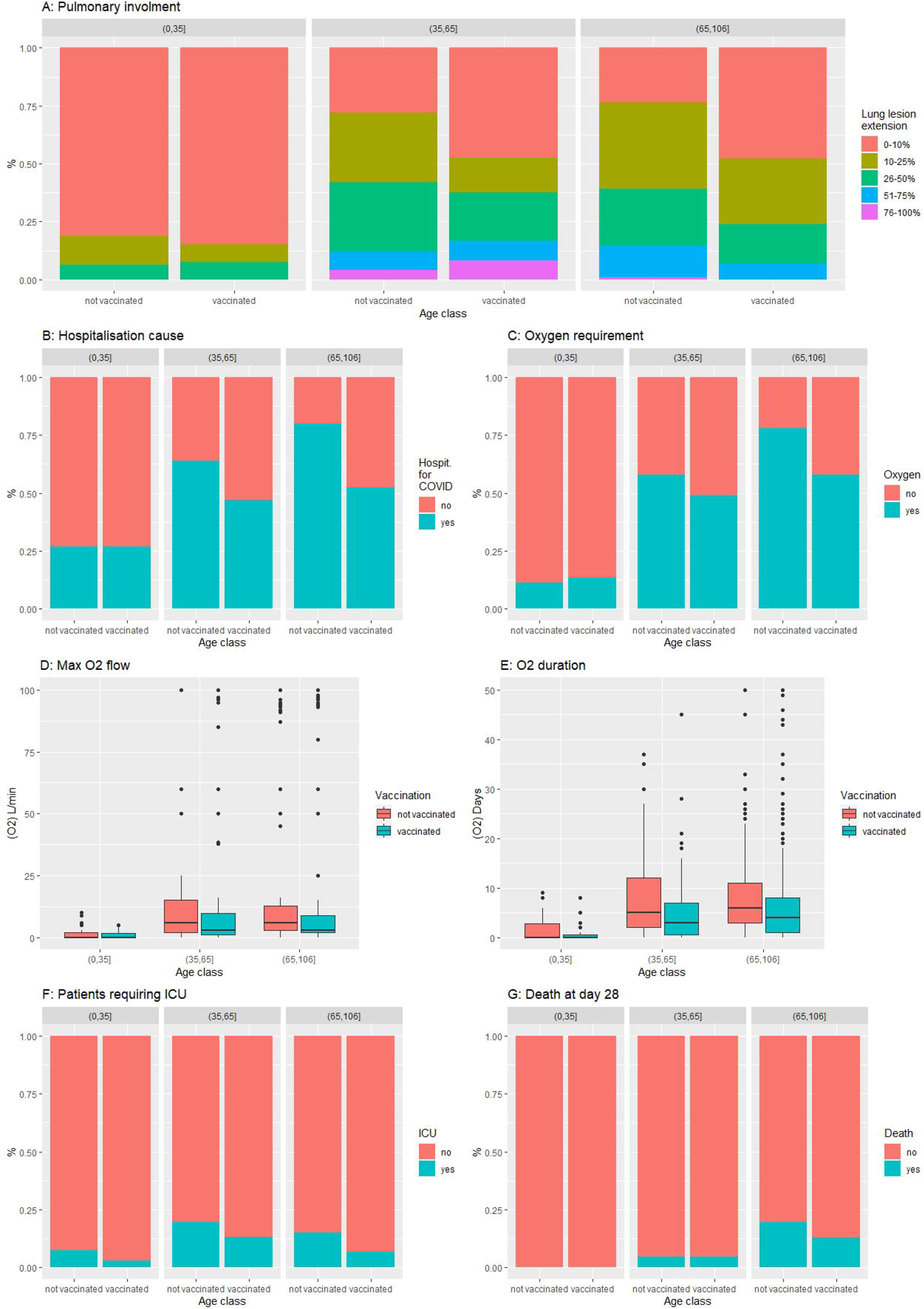
SARS-CoV-2 (Omicron variant) infection severity criteria according to vaccine status in the three age classes. A: Extension of lung lesions on CT scanner; B: hospitalisation cause; C: Proportion of patients requiring oxygen; D: Maximal oxygen flow; E: Duration of oxygen therapy; F: Proportion of patients requiring ICU and G: Proportion of death at day 28.

### Factors associated with severity

Three outcomes were considered: the need to receive oxygen, the need to be admitted to an ICU, and death at day 28. In bivariate analysis, older age, male gender and most comorbidities (as well as having at least 1 comorbidity, or a higher number of comorbidities) were significantly associated with one or several negative outcomes. Conversely, vaccination was protective against these three outcomes (Table 3).

In multivariate analysis, the protective effect of vaccination persisted with regard to need for oxygen, need for ICU admission, and risk of death, and it attenuated and practically discarded the negative impact of many comorbidities (Table 4). Older age, solid or haematological cancer and kidney failure remained independent risk factors for a more severe outcome, notably death. Pulmonary diseases tended to negatively impact the outcome but only regarding need for oxygen was the impact significant. A past history of SARS-CoV-2 infection tended to be protective against death and the need for ICU but with large confidence intervals that excluded significance.

### Influence of the variant of concern (Delta vs. Omicron)

Patients infected by the Omicron VOC presented less severe infection, either vaccinated or unvaccinated, compared to patients infected by the Delta VOC.

In bivariate analysis (Table 5 and 6), whether vaccinated or not, Omicron-infected patients were significantly (or at least presented a trend) less hospitalised for COVID-19 (significantly among those vaccinated), had lower median CRP (significantly among those vaccinated), had less extended lung lesion on CT scan (significantly among those unvaccinated), less frequently required oxygen supplementation, high-flow oxygen therapy, steroid therapy or tocilizumab (significantly among both vaccinated and unvaccinated), less frequently required to be admitted in ICU (both vaccinated and unvaccinated), and had lower 28-day mortality (among those vaccinated).

**Table 5:** Comparison of the Omicron infection with the Delta infection according to vaccination status: Demographics, underlying diseases and presentation.

|  | Vaccinated (N=1220) |  |  | Unvaccinated (N=950) |  |  |
| --- | --- | --- | --- | --- | --- | --- |
|  | Omicron<br>(n=746) | Delta<br>(n=474) | p-value | Omicron<br>(n=469) | Delta (n=481) | p-value |
| Age (years) n (%) |  |  | 0.031 |  |  | <0.001 |
| <35y | 67 (9.0) | 27 (5.7) |  | 54 (11.6) | 97 (20.2) |  |
| 35-65y | 198 (26.6) | 111 (23.4) |  | 128 (27.4) | 225 (46.8) |  |
| >65y | 481 (64.5) | 336 (70.9) |  | 287 (61.2) | 159 (33.1) |  |
| Age (years) | 74.0 [57.3;<br>85.0] | 75.0 [63.3;<br>84.0] | 0.012 | 71.0 [55.0;<br>83.0] | 55.0 [38.0;<br>73.0] | <0.001 |
| Gender: men n (%) | 406 (54.4) | 261 (55.1) | 0.873 | 217 (46.3) | 223 (46.4) | 1.000 |
| Body Mass Index<br>(kg/m<sup>2</sup>) | 25.9 [22.2;<br>29.4] | 25.8 [22.3;<br>31.1] | 0.036 | 26.1 [22.6;<br>29.2] | 27.2 [24.2;<br>31.2] | <0.001 |
| Overweight<br>(BMI>25) n (%) | 221 (56.5) | 192 (56.6) | 1.000 | 134 (57.8) | 222 (69.4) | 0.006 |
| Pre-existing<br>comorbidities n (%) |  |  |  |  |  |  |
| Cardiac failure | 121 (23.6) | 50 (13.7) | <0.001 | 65 (18.3) | 29 (8.0) | <0.001 |
| Respiratory disease | 103 (13.8) | 91 (20.7) | 0.002 | 47 (10.0) | 54 (12.6) | 0.256 |
| Oxygen at home | 15 (2.9) | 14 (4.0) | 0.515 | 3 (0.8) | 5 (1.4) | 0.501 |
| Kidney failure<br>(GFR<30) | 88 (12.0) | 51 (15.2) | 0.182 | 31 (6.8) | 10 (3.2) | 0.041 |
| Diabetes | 184 (24.7) | 127 (28.7) | 0.141 | 106 (22.6) | 76 (17.8) | 0.086 |
| Hypertension | 261 (50.8) | 244 (55.1) | 0.206 | 183 (51.5) | 123 (28.7) | <0.001 |
| Solid cancer < 3<br>months | 22 (3.0) | 17 (4.8) | 0.170 | 9 (1.9) | 8 (2.2) | 0.945 |
| Hematologic<br>malignancy | 42 (5.6) | 25 (7.1) | 0.428 | 10 (2.1) | 6 (1.7) | 0.869 |
| Immunodepression | 77 (15.0) | 74 (20.3) | 0.048 | 33 (9.3) | 21 (5.8) | 0.110 |
| At least one<br>comorbidity among<br>those above | 523 (70.1) | 369 (90.4) | <0.001 | 330 (70.4) | 306 (86.9) | <0.001 |
| Number of<br>comorbidities | 1.0 (0.0; 2.0] | 2.0 [1.0; 3.0] | <0.001 | 1.0 [0.0; 2.0] | 1.0 [0.0; 2.0] | 0.010 |
| Tobacco, active or discontinued < 3 years, n (%) | 44 (11.9) | 36 (10.3) | 0.580 | 38 (15.4) | 49 (14.3) | 0.800 |
| Previous SARS-CoV-2 infection n (%) | 12 (1.9) | 19 (5.4) | 0.005 | 10 (2.4) | 12 (3.4) | 0.533 |
| Time from a previous infection (days) | 319.0 [43.0; 344.0] | 226.0 [176.5; 299.0] | 0.688 | 215.0 [97.3; 389.5] | 69 [43.3; 244.3] | 0.113 |
| Time from last vaccine dose (days)* | 113.0 [64.0; 178.2] | 125.0 [82.0; 166.5] | 0.716 | - | - | - |
| Admission for COVID-19 | 359 (48.7) | 281 (59.3) | <0.001 | 317 (69.5) | 361 (75.1) | 0.069 |
| Ct value (lowest value if many) | 21.4 [18.2; 27.0] | 26.0 [18.0; 34.0] | <0.001 | 22.1 [19.0; 27.3] | 26.0 [21.0; 33.0] | <0.001 |
| CRP (highest value) (mg/L) | 58.0 [21.5; 117.5] | 87.5 [31.3; 152.8] | <0.001 | 66.0 [26.0; 126.3] | 86.0 [34.0; 145.0] | 0.073 |
| Proportion of lung involved on CT scan n (%) |  |  | 0.134 |  |  | <0.001 |
| 0-10% | 90 (50.3) | 67 (36.8) |  | 51 (30.4) | 31 (13.5) |  |
| 11-25% | 40 (22.3) | 50 (27.5) |  | 55 (32.7) | 57 (24.9) |  |
| 26-50% | 32 (17.9) | 39 (21.4) |  | 41 (24.4) | 75 (32.8) |  |
| 51-75% | 12 (6.7) | 18 (9.9) |  | 18 (10.7) | 49 (21.4) |  |
| >75% | 5 (2.8) | 8 (4.4) |  | 3 (1.8) | 17 (7.4) |  |

**Table 6:** Comparison of the Omicron infection with the Delta infection according to vaccinal status: Management and outcomes during hospital stay.

|  | Vaccinated (N=1220) |  |  | Unvaccinated (N=950) |  |  |
| --- | --- | --- | --- | --- | --- | --- |
|  | Omicron<br>(n=746) | Delta<br>(n=474) | p-value | Omicron<br>(n=469) | Delta (n=481) | p-value |
| Requirement for oxygen n (%) | 385 (51.6) | 229 (57.5) | 0.064 | 304 (64.8) | 270 (73.0) | 0.014 |
| Maximal oxygen therapy (L/min) | 3.0 [1.0; 9.0] | 3.0 [0.0; 8.8] | 0.140 | 6.0 [2.0; 15.0] | 6.0 [2.0; 50.0] | 0.153 |
| Duration of oxygen therapy (days) | 3.0 [0.5; 7.3] | 3.0 [0.0; 8.0] | 0.369 | 6.0 [2.0; 10.0] | 6.0 [2.0; 12.0] | 0.629 |
| High-flow oxygen n (%) | 32 (4.4) | 49 (13.6) | <0.001 | 56 (12.0) | 105 (29.4) | <0.001 |
| Steroid therapy |  |  |  |  |  |  |
| prescribed n (%) | 297 (40.0) | 174 (47.2) | 0.027 | 275 (58.8) | 250 (68.5) | 0.005 |
| dose median (IQR) | 7.5 [6.0; 10.7] | 6.0 [6.0; 7.5] | <0.001 | 8.0 [6.0; 12.0] | 6.0 [6.0; 9.0] | <0.001 |
| duration (days) | 5.0 [3.0; 10.0] | 9.0 [5.0; 10.0] | <0.001 | 8.0 [4.0; 10.0] | 10.0 [7.0; 10.0] | <0.001 |
| Tocilizumab n (%) | 20 (2.7) | 21 (6.3) | 0.007 | 29 (6.2) | 59 (18.3) | <0.001 |
| Monoclonal Ab n (%) | 30 (4.0) | 27 (7.3) | 0.030 | 19 (4.1) | 7 (1.9) | 0.122 |
| ICU admission n (%) | 60 (8.0) | 60 (16.2) | <0.001 | 72 (15.4) | 133 (36.0) | <0.001 |
| Invasive mechanical ventilation n (%) | 17 (2.3) | 25 (6.9) | <0.001 | 22 (4.7) | 59 (16.5) | <0.001 |
| Death at day 28 n (%) | 71 (9.5) | 74 (16.7) | <0.001 | 62 (13.2) | 53 (12.2) | 0.733 |
| Death at day 28 and/or ICU admission n, (%) | 113 (15.1) | 116 (30.7) | <0.001 | 116 (24.7) | 166 (44.6) | <0.001 |
| Length of stay | 7.0 [3.0; 14.0] | 9.0 [4.0; 16.0] | 0.003 | 8.0 [4.0; 13.0] | 7.0 [3.0; 15.0] | 0.866 |

In multivariate analysis (Table 7), Omicron infection, compared to Delta, was associated with less need for oxygen therapy, less ICU admission, and fewer deaths, while vaccination and a past history of SARS-CoV-2 infection were also protective. Only male gender, older age, kidney failure, immunosuppression and solid cancer were still risk factors for mortality. The interaction between immunosuppression and vaccination suggests that the protective effect of vaccination on the need for ICU is decreased among the immunosuppressed. The interaction between vaccination and a past history of SARS-CoV-2 infection suggests a lesser benefit of vaccination among patients having previously had SARS-CoV-2 infection.

**Table 7:** Multivariate analysis on the merged cohort (delta + omicron).

|  | Death on day 28 |  | ICU |  | Need for oxygen |  |
| --- | --- | --- | --- | --- | --- | --- |
|  | Best model | p-value | Best model | p-value | Best model | p-value |
| Variant: Omicron | 0.53 [0.37; 0.76] | 0.001 | 0.19 [0.12; 0.28] | <0.001 | 0.50 [0.38; 0.67] | <0.001 |
| Gender: Male | 1.66 [1.15; 2.41] | 0.007 | 1.37 [0.96; 1.95] | 0.084 | 1.65 [1.25; 2.18] | <0.001 |
| Age | 1.07 [1.05; 1.08] | <0.001 |  |  | 1.04 [1.03; 1.04] | <0.001 |
| Kidney failure | 2.32 [1.46; 3.67] | <0.001 | 0.41 [0.15; 0.93] | 0.050 |  |  |
| HTA | 1.28 [0.87; 1.90] | 0.211 |  |  |  |  |
| Immunosuppressed | 2.21 [1.30; 3.66] | 0.003 | 0.79 [0.31; 1.83] | 0.593 | 1.56 [1.05; 2.33] | 0.028 |
| Previous SARS-CoV-2 infection | 0.14 [0.01; 0.67] | 0.053 | 0.10 [0.01; 0.50] | 0.028 | 0.08 [0.02; 0.26] | <0.001 |
| Vaccination | 0.45 [0.30; 0.66] | <0.001 | 0.26 [0.17; 0.39] | <0.001 | 0.20 [0.14; 0.28] | <0.001 |
| Solid Cancer < 3 months | 5.61 [2.53; 12.57] | <0.001 | 0.41 [0.09; 1.31] | 0.174 |  |  |
| Overweight |  |  | 1.72 [1.10; 2.77] | 0.021 | 1.63 [1.17; 2.27] | 0.004 |
| At least one comorbidity |  |  | 1.68 [0.84; 3.45] | 0.146 | 1.61 [0.97; 2.67] | 0.064 |
| O2 treatment at home |  |  |  |  | 2.86 [1.05; 9.28] | 0.053 |
| Interaction vaccination x Immunosuppressed * |  |  | 2.95 [1.03; 8.87] | 0.047 |  |  |
| Interaction vaccination x Previous COVID-19* |  |  |  |  | 5.60 [1.21; 28.49] | 0.030 |

In the model for ICU, a significant and positive interaction was found between being vaccinated and being immunosuppressed, which means that the protective effect of vaccination on the risk of ICU admission decreases among immunosuppressed patients.
In the model for O<sub>2</sub> need, a significant and positive interaction was found between being vaccinated and having a previous SARS-CoV-2 infection, which means that effect of vaccination was decreased among patients having previously had SARS-CoV-2 infection. In addition, a
trend toward interaction between being vaccinated and being immunosuppressed was found, similarly to the ICU model, but was not significant ( $p=0.08$ ), which suggests an effect comparable to the ICU model.

## Discussion

Despite the fact that Omicron emerged as a new VOC at the end of 2021, our results show that vaccines designed in early 2020 still protect from severe forms. This protection persists even, if not mostly, in older patients, as well as patients with comorbidities. Our study also confirms that Omicron is less associated with severe forms than Delta, independently from the known risk factors and from vaccine status. Complementarily, vaccine-induced protection prevails, whether the VOC is a Delta or Omicron. However, greater age, kidney failure, and solid or haematological cancer are still associated with mortality, whereas vaccination can limit the influence of many risk factors.

The large cohort of 2167 patients obtained by merging our two cohorts highlights the fact that the VOC is a risk factor in itself, with a greater risk of severe form associated with Delta rather than Omicron, but also highlights the impact of past history of SARS-CoV-2 infection. Previous SARS-CoV-2 infection protected from the need for oxygen or ICU in the Delta cohort, but only with a non-significant trend toward lower mortality (Epaulard et al. 2022). A similar but likewise non-significant trend (for the three outcomes) was observed in the current study on Omicron. That said, the whole cohort provided sufficient power to show a significant influence of a past SARS-CoV-2 infection regarding mortality, need for ICU or need for oxygen.

Our results regarding the positive impact of vaccination on the SARS-CoV-2 infection are in line both with previous studies on previous variants (particularly Delta (Epaulard et al. 2022; Katikireddi et al. 2022; Harder et al. 2021)) and Omicron (Katikireddi et al. 2022; Brosh-Nissimov et al. 2022). Some of these studies evidenced a marked decrease of vaccine efficacy toward SARS-CoV-2 infection, but conserved efficacy regarding severe infection and death. Most of the studies included patients even if they were not hospitalised; the fact that we observed this protection even among patients hospitalised for Covid-19 (compared to those with an incidental SARS-CoV-2 infection with no clinical Covid-19) is a strong argument confirming the benefits of the vaccine.

Our observation regarding the lower risk of severe forms with Omicron infection compared with Delta is in accordance with the current literature. Indeed, previous studies concluded that Omicron was associated with less severe infections (Nyberg et al. 2022; Kahn et al. 2022). However, these studies were carried out at the populational level, where a large number of asymptomatic or mild cases could have gone undetected, thereby diluting the power to detect severe cases. Moreover, Delta and Omicron waves were not contemporaneous, and have spread in potentially very different populations in terms of vaccine coverage, past history of SARS-CoV-2 infection, etc. Such differences in the populations could generate differences misattributed to the virus. However, with a multivariate analysis applied to two cohorts with the same design, our study was able to decouple the effect of Omicron from vaccination, and consequently confirmed that it produces less severe forms than Delta. Nonetheless, the lesser proportion of severe forms with Omicron compared to Delta is not as marked among older people compared to the general population (Auvigne et al. 2022). It could be related to the difference of age between the Delta and the Omicron cohort, but only partially as the multivariate analysis showed an effect independent between Age and VOC. Consequently, severe forms do exist with Omicron and are not negligible. Consequently, the benefit of vaccination even among older patients and patients with comorbidities is an important rationale for vaccine campaigns. Studies showing the benefit of vaccination among specific subpopulations at risk also question the necessity of booster doses. As an example, studies in hemodialysis patients have confirmed a vaccine-induced protection proportional to the number of doses (Cheng et al. 2022; Espi et al. 2022).

Our study presents some limitations, in particular missing data, mostly due to its retrospective design; for that reason, some items were not taken into consideration in the analysis such as vaccine type. Moreover, we included patients from four countries, and our results may have been different if more countries (either European or not) had been represented. In addition, we did not perform anti-Spike serology at admission; anti-Spike antibody level has been shown to be associated with prognosis (Plūme et al. 2022; Sanghavi et al. 2022), and it might have facilitated understanding of the severity of SARS-CoV-2 infection in some vaccinated subjects, in particular among immunosuppressed subjects who could present lesser response to vaccines. In addition, availability of antiviral treatment was not comparable between the Delta and the Omicron wave, which could have had an impact on mortality.

The main strength of our study is the large number of highly phenotyped patients, from several centres for several countries, making our conclusions probably valid for most populations. In particular, vaccine status was individually checked, ensuring accurate classification of patients. In addition, the inclusion period we chose corresponds to a phase of homogeneous circulation of a unique variant, ensuring more precision to our conclusions. Lastly, the fact that the data collection was aligned with the Delta study performed in 2021 enabled us to conduct with high validity the comparison between the two variants.

Our study has several implications. The fact that despite the immune escape by Omicron, vaccination with the currently available vaccines still retains efficacy against severe infections, should be remembered and widely publicised, to help the population at risk of severe COVID-19 and their healthcare professionals to receive an additional dose.

Interestingly, if Omicron infection is less associated with severe COVID-19, it was also shown to be more contagious than Delta infection (Syed et al. 2022; Cui et al. 2022); this explains why many deaths were still observed during the BA-1 and BA-2 waves, the low frequency of severe form being counterbalanced by the massive number of infections. This observation undermines the statements describing Omicron as a step of SARS-CoV-2 toward a more “banal” coronavirus (Balloux et al. 2022; Callaway 2021; Gudina, Ali, et Froeschl 2022), such as 229E, NL63, OC43 and HKU1, and is an argument to maintain high vaccine coverage. Moreover, divalent mRNA vaccines have received market authorisation by the European medicines agency (EMA 2022 for Wuhan SARS-CoV-2 and Omicron BA.1 SARS-CoV-2 on 01/09/2022), and by the Food and Drug Administration (FDA) of the USA (FDA 2022 Wuhan SARS-CoV-2 and Omicron BA.4/5 on 31/08/2022); our results suggest that patients with risk factors for severe Covid-19 should not wait for such vaccines before receiving a boosting injection (such an injection being recommended 3 to 6 months after the previous one), as currently available vaccines, targeting the Wuhan SARS-CoV-2 viral sequence, still retain efficacy.

During the second quarter of 2022, new Omicron subvariants, BA.4 and BA.5, progressively became predominant in Europe. Studies showed a decrease in the neutralisation activities of antibodies elicited by vaccination (Wang et al. 2022) and of therapeutic monoclonal antibodies (Takashita et al. 2022) toward them. New studies are needed to determine the extent to which vaccination status, the number of vaccine doses and the timing of the last injection bring protection against severe infections, in addition to the impact of the divalent vaccines.

## Data Availability

All data produced in the present study are available upon reasonable request to the authors

## Acknowledgements

Authors are indebted to Sami Kinikli and Metin Ozsoy for data capture, and to Jeffrey Arsham for English editing.

## Access to data

Anonymized data and analysi files are available on request to the corresponding author.

## Contribution

Conceptualization: GB, OE

Methodology: GB, OE

Software: GB, OE

Validation : SA, NT, JFF, JM, ML, PV, CJ, RC

Formal Analysis: GB, OE

Investigation : GB, SA, NT, JM, JFF, SB, ML, PV, RC, MD, CJ, OE

Resources: not applicable

Data curation: All authors.

Writing – Original Draft: GB, OE

Writing – Review & Editing: SA, ML, NT, PV, JFF, JM, SB, CJ

Visualization: GB, OE

Supervision: GB, OE

Administration: GB, OE

Funding acquisition: not applicable

