## Supplementary material for "Impact of vaccination on the presence and severity of symptoms of hospitalised patients with an infection by the Omicron variant (B.1.1.529) of the SARS-CoV-2 (subvariant BA.1)"

Appendix:

The following data were collected: year of birth, gender, and pre-existing comorbidities (respiratory disease, cardiac failure, arterial hypertension, solid tumour, haematological neoplasia, and kidney failure, which was determined by the Cockcroft formula based on data collected at hospital admission); COVID-19 vaccination history; clinical data at admission and minimum cycle threshold (Ct) PCR level (the number of amplification cycles required for the signal to cross the threshold; Ct number is inversely proportional to the amount of target nucleic acid in the sample); maximum C-reactive protein plasmatic level; proportion of lung involved on CT scan (if performed); need for oxygen treatment with duration and maximal flow; need for high-flow oxygen therapy; need for ICU admission; need for invasive mechanical ventilation; prescription of steroid, tocilizumab, and/or monoclonal antibodies; and mortality at day 28.

Supplementary table 1: Multivariate analysis on vaccinated (Delta and Omicron) patients only.

|  | **Death on day 28** | | **ICU** | | **Need for oxygen** | |
| --- | --- | --- | --- | --- | --- | --- |
|  | **Best model** | **p-value** | **Best model** | **p-value** | **Best model** | **p-value** |
| Variant: Omicron | 0.48 [0.27; 0.84] | 0.012 | 0.22 [0.01; 0.29] | <0.001 | 0.78 [0.54; 1.10] | 0.157 |
| Gender: Male | 1.81[1.02; 3.32] | 0.047 |  |  | 1.27 [0.89; 1.80] | 0.192 |
| Age | 1.05 [1.03; 1.08] | <0.001 | 0.99 [0.97; 1.00] | 0.106 | 1.02 [1.01; 1.04] | <0.001 |
| Kidney failure | 2.96 [1.55; 5.55] | 0.001 | 0.49 [0.16; 1.25] | 0.170 |  |  |
| BMI | 1.04 [0.99; 1.08] | 0.104 | 1.07 [1.03; 1.11] | <0.001 |  |  |
| Solid Cancer < 3M | 6.40 [2.21; 18.35] | 0.001 |  |  |  |  |
| Last vaccine delay (days) | 1.00 [1.00; 1.01] | 0.057 |  |  | 1.00 [1.00; 1.00] | 0.067 |
| Cardiac failure |  |  | 2.11 [0.98; 4.40] | 0.050 | 1.47 [0.93; 2.35] | 0.104 |
| Immunosuppressed | 2.09 [1.04; 4.10] | 0.033 | 2.17 [1.12; 4.09] | 0.018 |  |  |
| Previous COVID |  |  | 0.24 [0.01; 1.23] | 0.172 | 0.41 [0.15; 1.00] | 0.059 |
| Pulmonary diseases |  |  | 1.77 [0.93; 3.32] | 0.078 |  |  |
| Tobacco |  |  |  |  | 0.63 [0.35; 1.14] | 0.129 |
| O2 at home |  |  |  |  | 2.34 [0.81; 8.44] | 0.145 |
| At least one comorbidity |  |  |  |  | 1.99 [1.15; 3.55] | 0.016 |

Supplementary table 1: Multivariate analysis on patients hospitalised for COVID only.

|  | **Death on day 28** | | **ICU** | | **Need for oxygen** | |
| --- | --- | --- | --- | --- | --- | --- |
|  | **Best model** | **p-value** | **Best model** | **p-value** | **Best model** | **p-value** |
| Variant Omicron | 0.28 [0.16; 0.48] | <0.001 | 0.27 [0.24; 0.69] | <0.001 | 0.34 [0.23; 0.51] | <0.001 |
| Gender: Male | 1.99 [1.16; 3.50] | 0.015 |  |  | 1.49 [1.04; 2.13] | 0.029 |
| Age | 1.06 [1.04; 1.09] | <0.001 |  |  | 1.03 [1.02; 1.04] | <0.001 |
| HTA | 1.96 [1.07; 3.70] | 0.032 |  |  |  |  |
| BMI | 1.03 [0.98; 1.07] | 0.231 |  |  |  |  |
| Kidney failure | 3.01 [1.50; 5.92] | 0.002 | 0.98 [0.57; 1.65] | 0.955 |  |  |
| Immunosuppressed | 2.11 [1.07; 4.04] | 0.027 |  |  |  |  |
| Solid cancer < 3 months | 5.40 [1.68; 17.46] | 0.004 |  |  |  |  |
| Vaccination |  |  | 0.45 [0.33; 0.62] | <0.001 | 0.51 [0.36; 0.74] | <0.001 |
| At least one comorbidity |  |  | 2.53 [1.56; 4.28] | <0.001 | 1.68 [1.04; 2.67] | 0.032 |
| O2 at home |  |  |  |  | 3.E6 [0.00; 5.3E59] | 0.973 |
